# Epidemiologic Assessment of Pediatric Inflammatory Bowel Disease Presentation in NYC During COVID-19

**DOI:** 10.1101/2022.03.23.22272167

**Authors:** Janet Rosenbaum, Kenny Castro Ochoa, Faria Hasan, Alexa Goldfarb, Vivian Tang, Gitit Tomer, Thomas Wallach

**Author notes:** **Address correspondence to:** Thomas Wallach MD, Division of Pediatric Gastroenterology, 450 Clarkson Ave, MSC 49. Brooklyn, NY, 11203. **Funding/Support:** No funding was secured for this study. **Role of Funder/Sponsor (if any):** none. **Contributors’ Statement Page** Dr. Wallach conceptualized the study and organized the consortium of institutions. Dr. Rosenbaum and Dr. Wallach designed the study, data collection instrument, and analytical framework. Drs Castro Ochoa, Tang, Tomer, Hasan, and Goldfarb collected data, organized local IRB approval at their respective institutions, and edited the manuscript. Dr. Rosenbaum harmonized, cleaned, and analyzed the data. Dr. Castro Ochoa, Dr. Rosenbaum, and Dr. Wallach wrote the manuscript. All authors reviewed, revised, and approved the final manuscript as submitted and agree to be accountable for all aspects of the work.

## Abstract

SARS-nCoV2 may have increased capacity to generate autoimmune disease; multiple reports suggest increased risk of Type 1 Diabetes, and case reports suggest other autoimmune linkages. Inflammatory Bowel Disease (IBD) pathogenesis appears to be a mix of genetic susceptibility, microbial populations, and immune triggers such as infections. Given the perceived role of infection in pathogenesis, decreased incidence of all infections during the pandemic secondary to non-pharmaceutical interventions should decrease IBD incidence rates. The aim of this study was to evaluate the association between the Covid-19 pandemic and IBD presentation in NYC using data from new diagnoses at a consortium of institutions.

Using EMR systems all diagnoses at 4 collaborating institutions were retrieved from 2015-2021. We fit time series model (ARIMA) to the quarterly number of cases of each disease for January 2016-March 2020 and forecast the subsequent 21 months. We not only did not observe a decline in pediatric IBD secondary to absent viral illness but noted a statistically significant increase in Crohn’s Disease approximately 6 months after the initial 2020 COVID wave in NYC, and trends suggesting increases overall in IBD diagnoses above the existing trend towards increased disease presentation that pre-dated the pandemic. This data suggests that there may be a linkage between SARS-nCoV2 infection rates and subsequent pediatric IBD presentation, warranting further evaluation in the aftermath of the Omicron wave.

## Introduction

The SARS-nCoV2 pandemic has created multiple challenges, but emerging data suggests additional possible long-term complications in the form of increased presentation of autoimmune diseases. Multiple Analytic studies have demonstrated Type 1 Diabetes diagnoses linked with SARS-nCov2 emergence, and case series suggest other autoimmune linkages^1-3^. Inflammatory Bowel Disease (IBD) is an autoimmune condition typically presenting in late childhood and young adulthood, with pathogenesis thought to be a mix of genetic susceptibility, microbial populations, and immune triggers such as infections^4^. Given the hypothesized role of infection in pathogenesis, decreased incidence of all infections during the pandemic secondary to non-pharmaceutical interventions (NPI) would likely be expected to generate a decrease in IBD incidence^4, 5^. As the first US locus of a major SARS-nCoV2 outbreak, New York City represents a unique opportunity to identify any signal of altered IBD presentation secondary to infection and the NPI implemented to manage the pandemic. This study will evaluate any association between the Covid-19 pandemic and new IBD diagnoses in NYC using data from a consortium of institutions.

## Methods

### Data

After institutional review board approval of respective institutions, we extracted date of diagnosis for Crohn’s Disease (CD) (n=349) and Ulcerative Colitis (UC) (n=145) patients presenting in pediatric GI clinic diagnosed 2016-2021 from electronic health records (EHRs).

### Measures

Date of diagnosis was coded as year and month of diagnosis with the day removed for identifiability reasons. We measured the following demographic variables: year and month of birth and EHR-recorded sex (always or almost always sex assigned at birth). Age at diagnosis was measured in years; was at median 14.0 years (range 2-21 years) and was available for all patients. For two institutions, we were able to extract race, ethnicity, and zip code. Gender was a free entry field, and only male and female patients were present in the data; 44.0% were female and 56.0% were male.

Race and ethnicity were measured by a free-entry field, and race was available for 30% of the participants and ethnicity for 44% of participants. Race was coded as White for free entry fields of White, and coded Black for free entry fields including Black, African American, and the typographical error “Afriac-American” [sic]. Race was coded as missing for declined, unavailable, or blank. Ethnicity was coded as Hispanic for free entry fields including Dominican, Mexican, Hispanic, Latino unspecified, Chicano/a, Puerto Rican, Salvadoran, South American, Spaniard, and Spanish. Participant ethnicity was coded as missing for entries of Declined, blanks, not applicable, not known, unknown n/a, unavailable.

An individual-level association between covid-19 infection and IBD would give the strongest inference, so we gathered data about past Covid-19 infection, measured as EMR record of either a positive-Covid-19 antibody test or positive Covid-19 PCR test. However, only one patient tested positive for antibodies and only one patient tested positive by PCR, so these measures were not used. Children were under-tested (cite) and tests may not appear in their EHR where they received IBD diagnosis because they may have been tested at school, public hospital system, urgent care clinics, or health systems that do not contribute to these EHR systems.

### Analysis

Due to potentially different seasonal variation and time to presentation, we analyzed CD and UC separately and excluded 22 cases of indeterminate IBD.

Using April 2020 as the first month of the covid-19 pandemic, we compared pre-pandemic versus pandemic periods with the Pearson chi-square test and pairwise proportion tests with Holm’s correction. We fit an autoregressive integrated moving average model (ARIMA) to the quarterly number of cases of each disease for January 2016-March 2020 and forecast the subsequent 21 months (April 2020-December 2021) with 80% and 95% prediction intervals using the forecast library.^5^ The model for ulcerative colitis was ARIMA(1,0,0) with a mean of 1.91 cases per month (se=0.28), lag-1 autocorrelation (AR1) of 0.30 (se=0.13), residual standard deviation of 2.00. The forecast varied only slightly across the interval. For ulcerative colitis, beginning in August 2020 an average of 1.90 cases were forecast with 80% prediction interval (0.00, 3.80) and 95% prediction interval (−1.00, 4.81).

The model for Crohn’s disease was ARIMA(0,0,0) (white noise) with mean of 4.65 cases per month (se = 0.37) and residual standard deviation of 7.07. Mean absolute scaled error is less than 1, which means the model is better than the naïve model (repeating previous year.) The forecast was uniform across the interval because the number of Crohn’s disease cases per month were white noise. For Crohn’s disease, an average of 4.65 cases was forecast with 80% prediction interval (1.24, 8.06) and 95% prediction interval (−0.57, 9.86).

We evaluated the robustness of results to monthly versus quarterly time period and in a larger year span, using 2011-2021 data from 3 contributing institutions (Appendix).

To show seasonal patterns in diagnoses, we plotted total cases by month with loess regression and 95% confidence intervals using ggplot2 stratified by pandemic versus pre-pandemic diagnosis.^6^ Residual plots showed no substantial departures from normality or evidence of heteroskedasticity. The Durbin-Watson statistic showed no evidence of autocorrelation for Crohn’s disease for either period or for Ulcerative Colitis for the SARS-nCoV2 pandemic period.

## Results

Patients diagnosed during the pandemic did not differ in demographic characteristics from patients diagnosed before the pandemic (**Table 1**). Downstate had a larger share of cases during the pandemic (9.6% of cases at the 4 institutions) than before the pandemic (4.3% of cases) and Maimonides had a smaller share of cases during the pandemic (10.2% of cases) than before the pandemic (17.5% of cases) (p=0.03); there were no significant differences between other institutions. Gender, age at diagnosis, diagnosis, county of residence, race, and ethnicity did not differ pre-pandemic versus post-pandemic.

**Table 1:** Demographics of participants (2016-2021): All percentages are column percentages. 90

|  | All participants | Jan. 2016 – Mar. 2020 | April 2020 – Dec. 2021 | P-value |
| --- | --- | --- | --- | --- |
| N | 516 | 349 | 167 |  |
| **Gender** |  |  |  | 0.6 |
| Male | 295 (57.2%) | 196 (56.2%) | 99 (59.3%) |  |
| Female | 221 (42.8%) | 153 (43.8%) | 67 (40.7%) |  |
| **Age at diagnosis** |  |  |  | 0.8 |
| 0-4 | 19 (3.7%) | 13 (3.7%) | 6 (3.6%) |  |
| 5-12 | 164 (31.8%) | 114 (32.7%) | 50 (29.9%) |  |
| 13-17 | 266 (51.6%) | 180 (51.6%) | 86 (51.5%) |  |
| 18-21 | 67 (13.0%) | 42 (12.0%) | 25 (15.0%) |  |
| **Institution** |  |  |  | 0.02 |
| Downstate | 31 (6.0%) | 15 (4.3%) | 16 (9.6%) |  |
| Maimonides | 78 (15.1%) | 61 (17.5%) | 17 (10.2%) |  |
| Montefiore | 236 (45.7%) | 155 (44.4%) | 81 (48.5%) |  |
| NYU | 171 (33.1%) | 118 (33.8%) | 53 (31.7%) |  |
| **Race** |  |  |  | 0.6 |
| White | 106 (20.5%) | 76 (21.8%) | 30 (18.0%) |  |
| Black | 45 (8.7%) | 29 (8.3%) | 16 (9.6%) |  |
| Other or not reported | 365 (70.7%) | 244 (69.9%) | 121 (72.5%) |  |
| **Hispanic ethnicity** |  |  |  | 0.6 |
| Not Hispanic | 121 (23.4%) | 83 (23.8%) | 38 (22.9%) |  |
| Hispanic | 110 (21.3%) | 70 (20.1%) | 40 (24.0%) |  |
| Not reported | 285 (55.2%) | 196 (56.2%) | 89 (53.3%) |  |
| **Year of diagnosis** |  |  |  | n.a. |
| 2016 | 67 (13.0%) | 67 (19.2%) | 0 (0%) |  |
| 2017 | 84 (16.3%) | 84 (24.1%) | 0 (0%) |  |
| 2018 | 80 (15.5%) | 80 (22.9%) | 0 (0%) |  |
| 2019 | 90 (17.4%) | 90 (25.7%) | 0 (0%) |  |
| 2020 | 96 (18.6%) | 28 (8.0%) | 68 (40.7%) |  |
| 2021 | 99 (19.2%) | 0 (0%) | 99 (59.3%) |  |
| **Diagnosis** |  |  |  | 0.7 |
| Crohn's Disease | 349 (67.6%) | 237 (67.9%) | 112 (67.1%) |  |
| Ulcerative Colitis | 145 (28.1%) | 98 (28.1%) | 47 (28.1%) |  |
| Undetermined | 20 (3.9%) | 12 (3.4%) | 8 (4.8%) |  |
| Not reported | 2 (0.4%) | 2 (0.6%) | 0 (0%) |  |
| **Location** |  |  |  | 0.4 |
| Manhattan | 31 (6.0%) | 20 (5.7%) | 11 (6.6%) |  |
| Brooklyn | 206 (39.9%) | 148 (42.4%) | 58 (34.7%) |  |
| Queens | 21 (4.1%) | 12 (3.4%) | 9 (5.4%) |  |
| Bronx | 151 (29.3%) | 100 (28.6%) | 51 (30.5%) |  |
| Staten Island | 11 (2.1%) | 6 (1.7%) | 5 (3.0%) |  |
| Westchester | 42 (8.1%) | 28 (8.0%) | 14 (8.4%) |  |
| Rockland | 30 (5.8%) | 18 (5.2%) | 12 (7.2%) |  |
| Long Island | 11 (2.1%) | 10 (2.9%) | 1 (0.6%) |  |
| Other NY or PA or CT or NJ | 5 (1.0%) | 2 (0.6%) | 3 (1.8%) |  |
| Not reported | 8 (1.6%) | 5 (1.4%) | 3 (1.8%) |  |

P-value from chi-square test of association between time period (pre-pandemic versus pandemic) and each variable.

We noted more CD diagnoses than expected in June and July during the pandemic compared with before the pandemic in the loess plots **[Figure 1]**.

**Figure 1.**
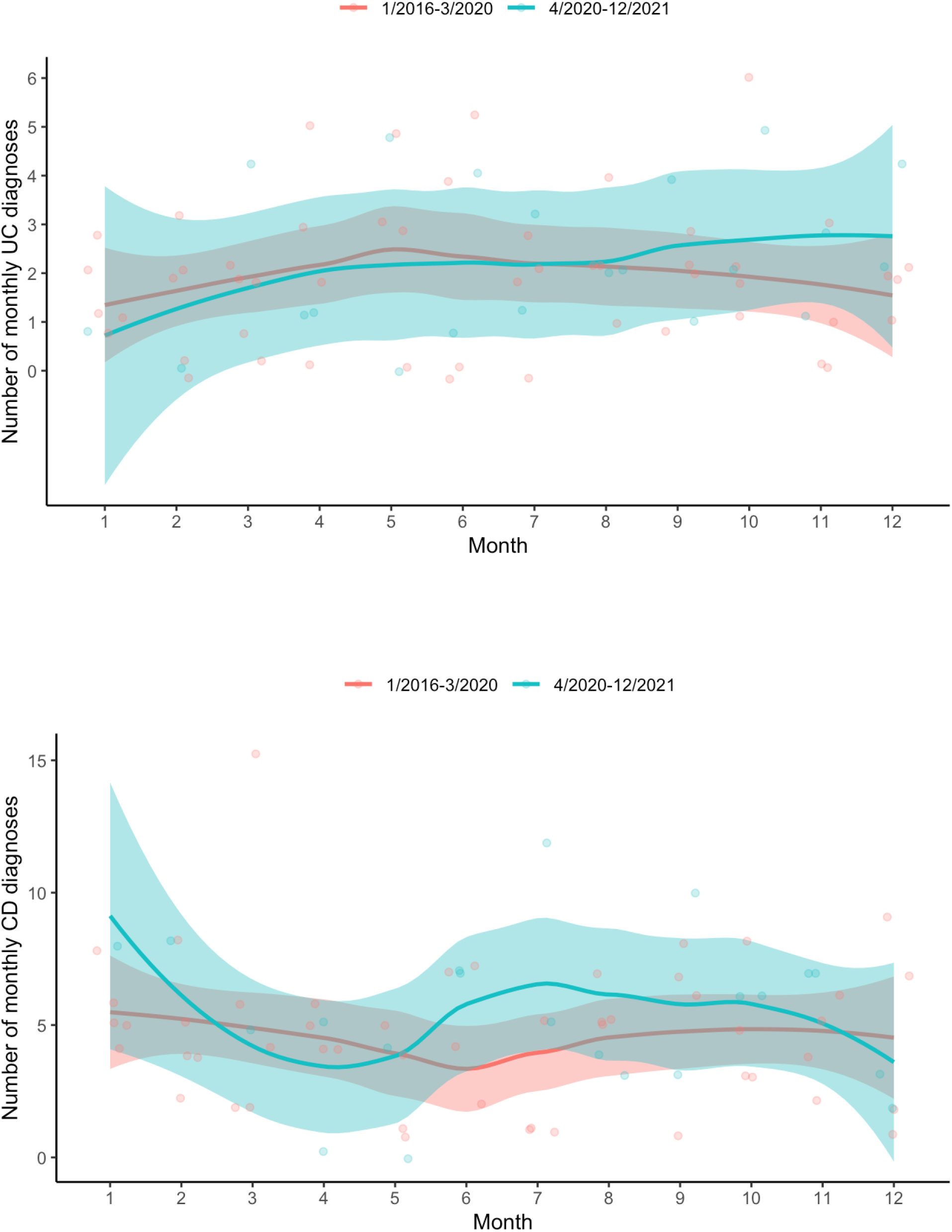
Loess plots of UC and CD presentation by month. A) Seasonal Loess plot of Ulcerative Colitis presentation by calendar month including all datapoints from 2016-2021. B) Seasonal Loess plot of Crohn’s Disease presentation by calendar month including all datapoints from 2016-2021

We noted more CD cases than forecast in the third quarter of 2020, which had 26 cases compared with upper limit of 95% prediction interval of 23.0 in ARIMA analysis **[Figure 2]**. We notice a trend towards elevation in UC diagnoses over time. Robustness checks using monthly data demonstrated similar rates exceeding 95% prediction in CD (July and September of 2020) and for UC in May and October of 2021, consistent with an elevation of mean monthly diagnoses.

**Figure 2.**
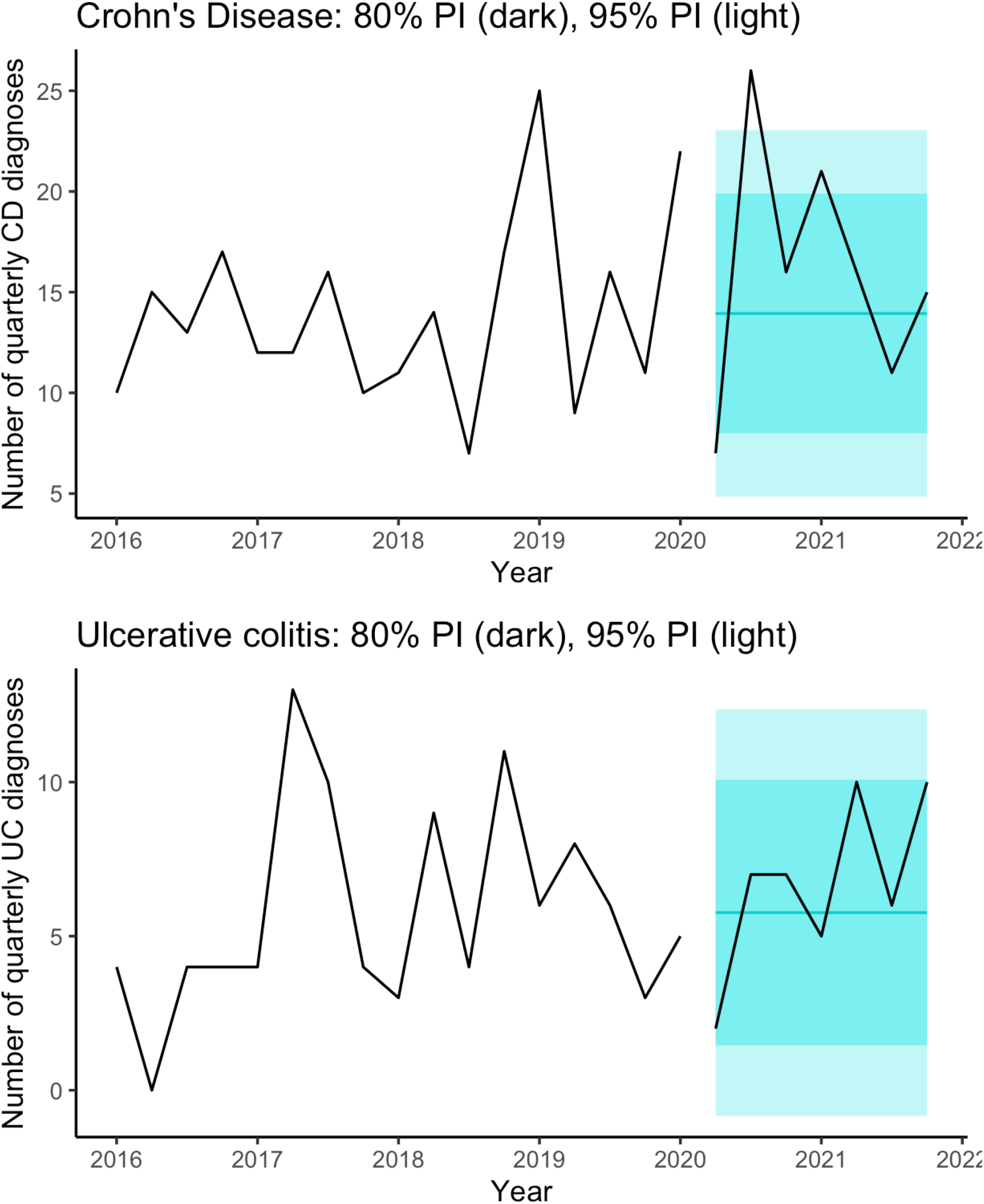
Time series (ARIMA) of UC and CD. A) ARIMA time series analysis of UC presentation by 3 month period from 2016-2021 with 80% and 95% prediction intervals.

As a first robustness check, we repeated the model with monthly data (2016-2021) **[Figure 3]**. The model for monthly ulcerative colitis was ARIMA (1,0,0) with a mean of 1.91 cases per month (se=0.28), lag-1 autocorrelation (AR1) of 0.30 (se=0.13), residual standard deviation of 2.00. The forecast varied only slightly across the interval. For ulcerative colitis, beginning in August 2020 an average of 1.90 cases were forecast with 80% prediction interval (0.00, 3.80) and 95% prediction interval (−1.00, 4.81). The model for Crohn’s disease was ARIMA (0,0,0) (white noise) with mean of 4.65 cases per month (se = 0.37) and residual standard deviation of 7.07. Mean absolute scaled error is less than 1, which means the model is better than the naïve model (repeating previous year.) The forecast was uniform across the interval because the number of Crohn’s disease cases per month were white noise. For Crohn’s disease, an average of 4.65 cases was forecast with 80% prediction interval (1.24, 8.06) and 95% prediction interval (−0.57, 9.86). We notice more ulcerative colitis cases than forecast in May 2021 and October 2021, which had 5 cases each compared with upper limit of 95% prediction interval 4.81, and more Crohn’s disease cases than forecast in July 2020 (12 cases), September 2020 (10 cases), compared with upper limit of 95% prediction interval 9.86.

**Figure 3:**
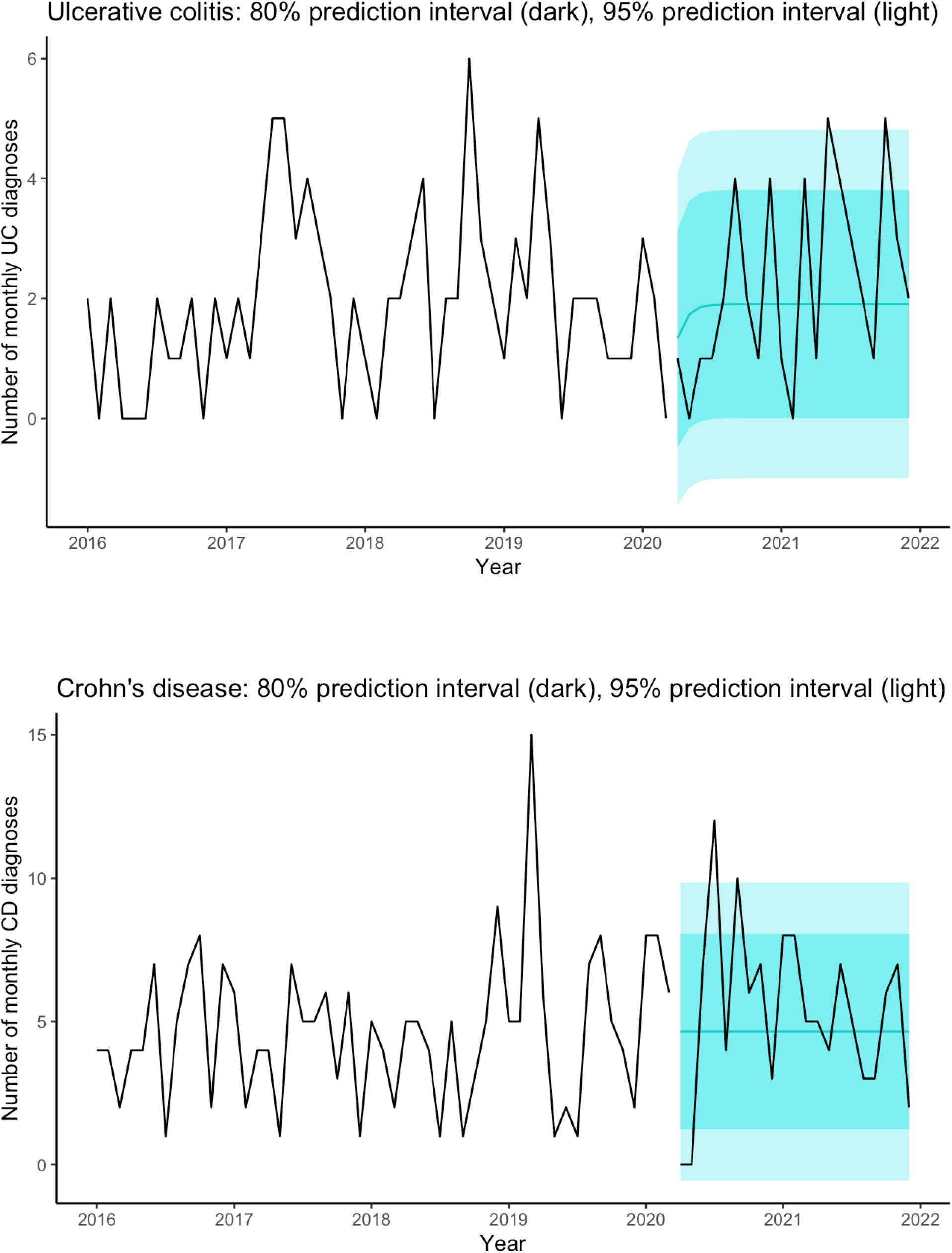
Time series with forecast: monthly ulcerative colitis (UC) and Crohn’s disease (CD) cases

As a second robustness check, we repeated the model with monthly and quarterly data from 2011-2021, which were available for 3 of the 4 institutions. **Table 2** compares the demographic data for these 3 institutions: as before, we notice that Downstate had more cases than usual during the pandemic than before the pandemic, and Maimonides had fewer, but there were no other differences; these three institutions do not report Hispanic ethnicity.

**Table 2:** Demographics of participants at 3 institutions, 2011-2021

|  | All participants | Jan. 2011 – Mar. 2020 | April 2020 – Dec. 2021 | P-value |
| --- | --- | --- | --- | --- |
| N | 466 | 380 | 86 |  |
| **Gender** |  |  |  | 0.6 |
| Male | 258 (55.4%) | 208 (54.7%) | 50 (58.1%) |  |
| Female | 208 (44.6%) | 172 (45.3%) | 36 (41.9%) |  |
| **Age at diagnosis** |  |  |  | 0.6 |
| 0-4 | 6 (1.3%) | 4 (1.1%) | 2 (2.3%) |  |
| 5-12 | 166 (35.6%) | 137 (36.1%) | 29 (33.7%) |  |
| 13-17 | 258 (55.4%) | 212 (55.8%) | 46 (53.5%) |  |
| 18-21 | 36 (7.7%) | 27 (7.1%) | 9 (10.5%) |  |
| **Institution** |  |  |  | 0.004 |
| Downstate | 42 (9.0%) | 26 (6.8%) | 16 (18.6%) |  |
| Maimonides | 110 (23.6%) | 93 (24.5%) | 17 (19.8%) |  |
| NYU | 314 (67.4%) | 261 (68.7%) | 53 (61.6%) |  |
| **Race** |  |  |  | 0.4 |
| White | 84 (20.5%) | 70 (18.4%) | 14 (16.3%) |  |
| Black | 7 (8.7%) | 7 (1.8%) | 0 |  |
| Other or not reported | 375 (70.7%) | 303 (79.7%) | 72 (83.7%) |  |
| **Hispanic ethnicity** |  |  |  | n.a. |
| Not Hispanic | 0 | 0 | 0 |  |
| Hispanic | 31 (6.7%) | 23 (6.1%) | 8 (9.3%) |  |
| Not reported | 435 (93.3%) | 357 (93.9%) | 78 (90.7%) |  |
| **Year of diagnosis** |  |  |  | n.a. |
| 2011 | 33 (7.1%) | 33 (8.7%) | 0 (0%) |  |
| 2012 | 27 (5.8%) | 27 (7.1%) | 0 (0%) |  |
| 2013 | 34 (7.3%) | 34 (8.9%) | 0 (0%) |  |
| 2014 | 36 (7.7%) | 36 (9.5%) | 0 (0%) |  |
| 2015 | 56 (12.0%) | 56 (14.7%) | 0 (0%) |  |
| 2016 | 39 (8.4%) | 39 (10.3%) | 0 (0%) |  |
| 2017 | 45 (9.7%) | 45 (11.8%) | 0 (0%) |  |
| 2018 | 43 (9.2%) | 43 (11.3%) | 0 (0%) |  |
| 2019 | 53 (11.4%) | 53 (13.9%) | 0 (0%) |  |
| 2020 | 50 (10.7%) | 14 (3.7%) | 36 (41.9%) |  |
| 2021 | 50 (10.7%) | 0 (0%) | 50 (58.1%) |  |
| **Diagnosis** |  |  |  | 0.3 |
| Crohn's Disease | 327 (70.2%) | 267 (70.3%) | 60 (69.8%) |  |
| Ulcerative Colitis | 126 (27.0%) | 104 (27.4%) | 22 (25.6%) |  |
| Undetermined | 10 (2.1%) | 6 (1.6%) | 4 (4.7%) |  |
| Not reported | 3 (0.6%) | 3 (0.8%) | 0 (0%) |  |
| **Location** |  |  |  | 0.2 |
| Manhattan | 49 (10.5%) | 38 (10.0%) | 11 (12.8%) |  |
| Brooklyn | 313 (67.2%) | 257 (67.6%) | 56 (65.1%) |  |
| Queens | 31 (6.7%) | 23 (6.1%) | 8 (9.3%) |  |
| Bronx | 6 (1.3%) | 4 (1.1%) | 2 (2.3%) |  |
| Staten Island | 13 (2.8%) | 9 (2.4%) | 4 (4.7%) |  |
| Westchester | 4 (0.9%) | 4 (1.1%) | 0 |  |
| Rockland | 14 (3.0%) | 10 (2.6%) | 4 (4.7%) |  |
| Long Island | 24 (5.2%) | 23 (6.1%) | 1 (1.2%) |  |
| Other NY or PA or CT or NJ | 5 (1.1%) | 5 (1.3%) | 0 |  |
| Not reported | 7 (1.5%) | 7 (1.8%) | 0 |  |

P-value from chi-square test of association between each variable and pre-pandemic versus post-pandemic with simulated p-value.

The pre-pandemic model for quarterly ulcerative colitis was ARIMA(0,0,0) **[Figure 4]** with a mean of 2.81 cases per quarter (se = 0.28) and residual standard deviation of 2.88. The pre-pandemic model for quarterly Crohn’s disease was ARIMA(0,0,0) with a mean of 7.22 cases per quarter (se = 0.54) and residual standard deviation of 11.2. Mean absolute scaled error for the pandemic data for UC and CD were 0.92 and 0.82, which are less than 1, which mean the models are better than the naïve model (repeating previous year.)

**Figure 4:**
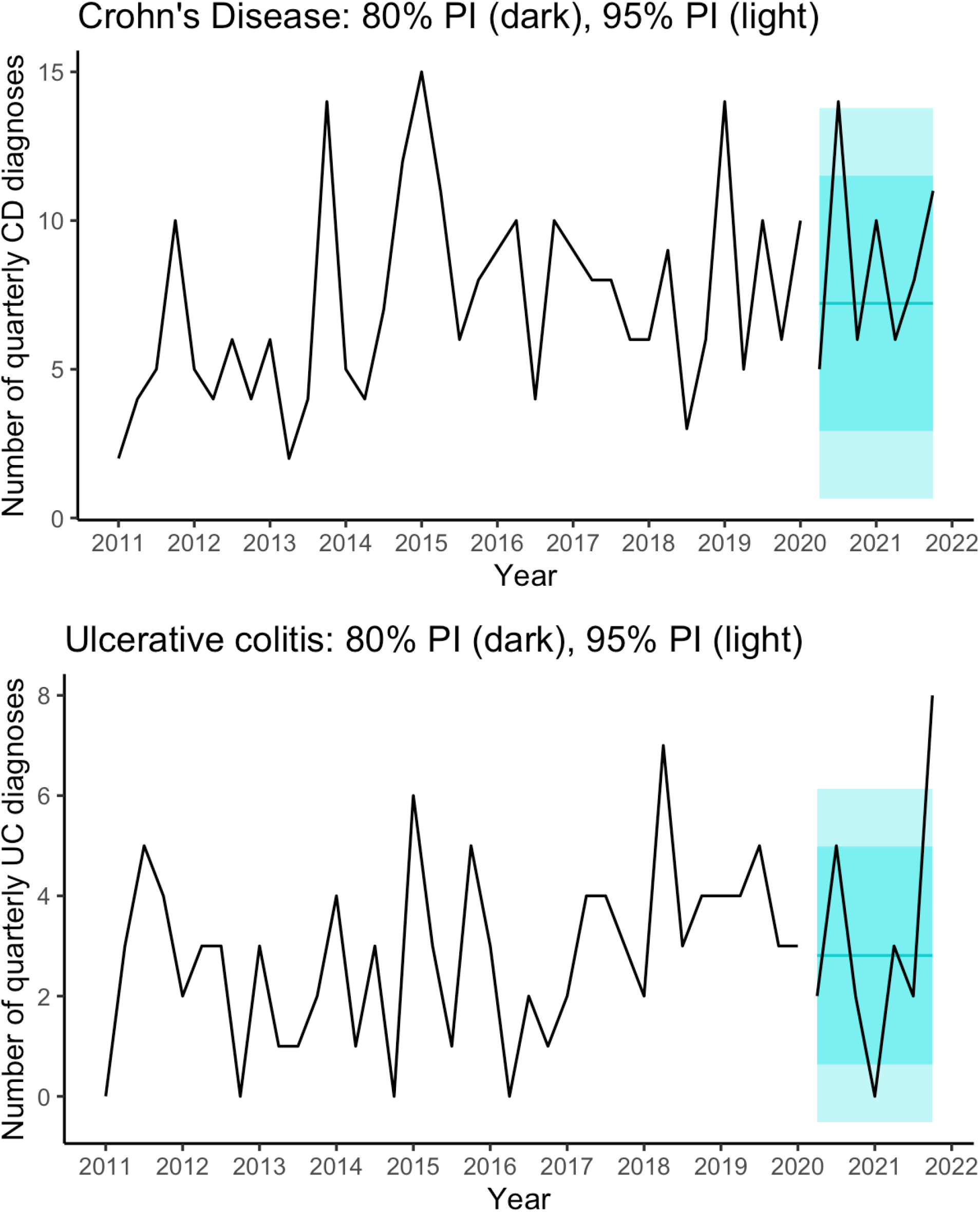
Time series with prediction intervals (PI) forecast: quarterly ulcerative colitis (UC) and Crohn’s disease (CD) cases, 2011-2021.

**Figure 5:**
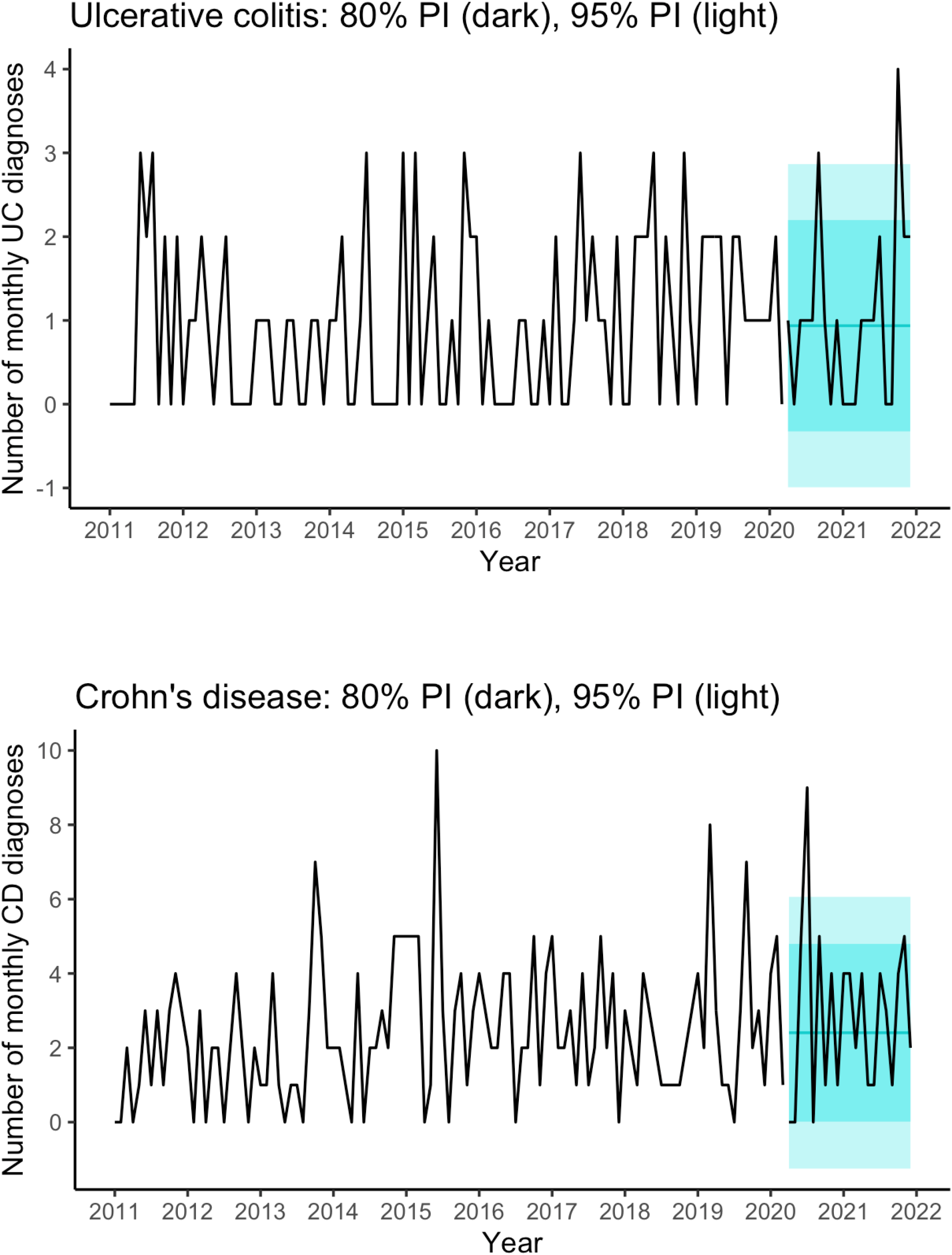
Time series with prediction intervals (PI) forecast for 3 institutions: monthly ulcerative colitis (UC) and Crohn’s disease (CD) cases.

With these three institutions, we notice more Crohn’s disease than forecast in the third quarter of 2020 which had 14 cases compared with upper limit of 95% prediction interval of 13.8, and more ulcerative colitis than forecast in the fourth quarter of 2021, which had 8 cases compared with upper limit of 95% prediction interval of 6.1. We notice a trend towards elevation in UC diagnoses over time.

## Discussion

Decreases in viral infection did not decrease CD and UC presentation rates, suggesting either diminished importance of viral infection as a pathogenic trigger of IBD or a strong capacity for SARS-nCoV2 to drive IBD pathogenesis. Second, we observe a significant increase in CD diagnoses with a possible temporal linkage of 4-6 months after pre-omicron peak SARS-nCoV2 infection in NYC in March/April 2020.

Our study has several limitations. IBD does not present with consistent patterns, resulting in wide variability in data. The 4-6 month lag between the city’s peak infection rate and CD is more consistent with a SARS-nCoV2 infection pathogenesis than with delayed presentation from cases during the lockdown period. Our data does not include all pediatric gastroenterology groups in NYC, which may create effects due to variations in patient choice of providers.

Our study suggests a risk that large increases of SARS-nCoV2 infection in pediatric patients may drive increases in IBD diagnoses, in particular CD. Given recent widespread infection during the omicron wave, this study highlights the necessity of follow-up investigation of changes in incidence using existing data infrastructure.

## Data Availability

All data produced in the present study are available upon reasonable request to the authors

## Abbreviations

(UC): Ulcerative Colitis
(CD): Crohn’s Disease
(IBD): Inflammatory Bowel Disease
(NYC): New York City

## Acknowledgments

We would like to acknowledge the contributions of the following individuals, Dalia Arostegui MD, Chika Oragui MD, Agata Mann MD, Jon Dooley MD, and Dr. Melanie Greifer MD who assisted in data collection, and Steven Schwarz MD who assisted in building our data gathering collaboration.

